# Targeting myeloperoxidase to reduce neuroinflammation in X-linked dystonia parkinsonism

**DOI:** 10.1101/2024.06.25.24309481

**Authors:** Tiziana Petrozziello, Negin J. Motlagh, Ranee Z.B. Monsanto, Dan Lei, Micaela G. Murcar, Ellen B. Penney, D. Cristopher Bragg, Cara Fernandez-Cerado, G. Paul Legarda, Michelle Sy, Edwin Muñoz, Mark C. Ang, Cid Czarina E. Diesta, Can Zhang, Rudolph E. Tanzi, Irfan A. Qureshi, John W. Chen, Ghazaleh Sadri-Vakili

**Affiliations:** Sean M. Healey & AMG Center for ALS at Mass General, Massachusetts General Hospital, Boston, MA 02129, USA; Institute for Innovation in Imaging, Department of Radiology, Massachusetts General Hospital, Boston, MA 02129, USA; Center for Systems Biology, Massachusetts General Hospital and Harvard Medical School, Boston, MA 02114, USA; Department of Neurology, Massachusetts General Hospital, Boston, MA 02129, USA; Sunshine Care Foundation, 5800 Roxas City, Capiz, Philippines; Department of Pathology, College of Medicine, University of the Philippines, Manila, Philippines; Department of Neurosciences, Makati Medical Center, Makati, Philippines; Biohaven Pharmaceuticals, Inc., New Haven, CT 06510, USA

**Keywords:** X-linked dystonia parkinsonism, myeloperoxidase, verdiperstat

## Abstract

Although the genetic locus of X-linked dystonia parkinsonism (XDP), a neurodegenerative disease endemic in the Philippines, is well-characterized, the exact molecular mechanisms leading to neuronal loss are not yet fully understood. Recently, we demonstrated a significant increase in astrogliosis and microgliosis together with an increase in myeloperoxidase (MPO) levels in XDP post-mortem prefrontal cortex (PFC), suggesting a role for neuroinflammation in XDP pathogenesis. Here, we demonstrated a significant increase in MPO activity in XDP PFC using a novel specific MPO-activatable fluorescent agent (MAFA). Additionally, we demonstrated a significant increase in reactive oxygen species (ROS) in XDP-derived fibroblasts as well as in SH-SY5Y cells treated with post-mortem XDP PFC, further supporting a role for MPO in XDP. To determine whether increases in MPO activity were linked to increases in ROS, MPO content was immuno-depleted from XDP PFC [MPO(-)], which resulted in a significant decrease in ROS in SH-SY5Y cells. Consistently, the treatment with verdiperstat, a potent and selective MPO inhibitor, significantly decreased ROS in both XDP-derived fibroblasts and XDP PFC-treated SH-SY5Y cells. Collectively, our results suggest that MPO inhibition mitigates oxidative stress and may provide a novel therapeutic strategy for XDP treatment.

**Highlights:**

□ MPO activity is increased in XDP post-mortem prefrontal cortex.
□ MPO activity is increased in cellular models of XDP.
□ MPO increases reactive oxygen species (ROS) in vitro.
□ Inhibiting MPO mitigates ROS in XDP.
□ The MPO inhibitor, verdiperstat, dampens ROS suggesting a potential therapeutic strategy for XDP.

## Introduction

X-linked dystonia parkinsonism (XDP) is an adult-onset, progressive, debilitating movement disorder that predominantly affects males with maternal ancestry originating from the island of Panay in the Philippines (Lee et al., 2011). All affected individuals reported to date share a common disease haplotype consisting of multiple sequence variants in and around the *TAF1* gene, which encodes for the TATA-binding protein-associated factor-1 (TAF1) (Nolte et al. 2003; Makino et al. 2007; Domingo et al., 2015; Aneichyk et al. 2018). All of these variants fall within non-coding gene regions, and only one has been directly linked to disease pathogenesis: a SINE-VNTR-Alu (SVA)-type retrotransposon insertion within intron 32 of *TAF1* (Makino et al., 2017; Aneichyk et al., 2018). This XDP-specific SVA element includes a 5’ tandem repeat, (CCCTCT)_n_, the length of which inversely correlates with age of disease onset in patients (Bragg et al., 2017; Westenberger et al. 2019), and this insertion disrupts transcription of *TAF1* in XDP cell models (Aneichyk et al. 2018; Al Ali et al., 2021). However, the exact molecular mechanisms by which these perturbations in *TAF1* lead to neuronal loss in XDP are still not completely elucidated.

Similar to other neurodegenerative diseases, such as Alzheimer’s disease (AD) (Thakur et al., 2023), Huntington’s disease (HD) (Saba et al., 2022), Parkinson’s disease (PD) (Araújo et al., 2022), and amyotrophic lateral sclerosis (ALS) (Obrador et al., 2020), a role for neuroinflammation in XDP pathogenesis has recently been suggested. Specifically, alterations in inflammatory pathways, including the dysregulation of NFκB signaling, hypersensitivity to TNF-α, and increases in several pro-inflammatory markers, were described in XDP-derived fibroblasts and neuronal stem cells (NSCs) (Vaine et al., 2017). Additionally, we have demonstrated an increase in astrogliosis and microgliosis as well as myeloperoxidase (MPO) levels in XDP post-mortem prefrontal cortex (PFC), further supporting a role for neuroinflammation in XDP pathogenesis (Petrozziello et al., 2020).

MPO is a member of the heme peroxidase family released from peripheral immune cells, such as neutrophils, macrophages, and monocytes (Nicholls et al., 2005), as well as from reactive astrocytes and activated microglia (Green et al., 2004; Choi et al., 2005; Gray et al., 2008; Forghani et al., 2015; Yu et al., 2016). Of note, prolonged increases in systemic levels of MPO have been linked to tissue damage during chronic inflammation, mostly due to degranulation and apoptosis of immune cells as well as prolonged increases in reactive oxygen species (ROS) (Babior, 2000; Shaeib et al., 2013; Parker et al., 2012; Hawkins and Davies, 2021). Therefore, we sought to determine whether increases in MPO contribute to neuroinflammation in XDP pathogenesis. First, we measured MPO activity in a large cohort of human post-mortem XDP PFC by using a novel MPO-activatable fluorescent molecular imaging probe (MAFA) (Motlagh et al., 2024). Next, we assessed the downstream consequences of increases in MPO in cellular models of XDP. Lastly, we sought to determine whether inhibiting MPO mitigates inflammation in XDP in vitro.

## Materials and Methods

### Human brain tissue samples

Post-mortem XDP PFC were provided by the Massachusetts General Hospital Collaborative Center for XDP (CCXDP) with approval of the Institutional Review Boards (IRB) at Massachusetts General Hospital (Boston, USA) and Makati Medical Center (Makati City, Philippines). Detailed methods on donor consent, tissue collection, and processing, as well as quality control metrics, have been previously reported (Fernandez-Cerado et al. 2021). Post-mortem control PFC were provided by the Massachusetts Alzheimer’s Disease Research Center (ADRC) with approval from the Massachusetts General Hospital IRB. This study included only control and XDP PFC derived from male subjects. The mean age was 78.9 years (SD = 13.03) for control subjects and 49.1 years (SD = 8.73) for XDP subjects. Age at disease onset for people who lived with XDP range was 27-59 years, disease duration range was 12-204 months, and repeat size within the SVA range was 34-52. Post-mortem interval (PMI) range was 16-36 h for control subjects and 6.36-35.52 h for XDP subjects.

### Ex vivo detection of MPO activity using MAFA

As a substrate for MPO, MPO can oxidize MAFA to form radicals that bind to nearby proteins containing phenolic or indolic moieties. Activated MAFA would then remain bound to tissues after washing, while inactivated agents would be washed away, enabling the identification of areas with elevated MPO activity (Motlagh et al., 2024). Fresh-frozen human brain sections (8 μm thickness) were fixed in 4% paraformaldehyde (PFA) for 5 min at room temperature (RT) and then incubated in blocking solution containing 1% fetal bovine serum (FBS), and 0.3% Triton X-100 (#T8787-50ml, MilliporeSigma, MA) at RT for 1 h. Following 3 washes in phosphate buffer saline (PBS), the sections were incubated with MAFA [1:300 of stock solution 10 mM dimethyl-sulfoxide (DMSO) and 1 mM of 3% hydrogen peroxide (H_2_O_2_)] for 30 min at RT. Lastly, the sections were mounted in an antifade mounting medium (#ZJ0808, Vectashield, Vector Laboratories, CA) containing the nuclear stain DAPI. Images were captured with a Nikon DS-Ri2 model microscope connect to Prime BSI Express. Intensity of fluorescence was measured by using ImageJ 1.53t (National Institute of Health, Bethesda, MD).

### Meso Scale Discovery (MSD) Assay

Cytokines levels were assessed using the human proinflammatory panel-1 10-plex kits to detect 10 cytokines, including interferon γ (INF-γ), interleukin (IL)-1β, IL-2, IL-4, IL-5, IL-6, IL-8, IL-10, IL-12p70, IL-13 and tumor necrosis factor-α (TNF-α). Specifically, cytokines levels were measured using an electrochemiluminescence-based multi-array method through the Quickplex SQ 120 system (Meso Scale Diagnostics LLC, MD) following previously reported methods (Raven, F., et al., 2017; Yang, H.S., et al., 2021). Briefly, samples or protein standards were incubated at 4°C overnight on a shaker. Next, samples and protein standards were washed and incubated for 2 h with the detection antibodies provided by the kit. Lastly, the electrochemiluminescence signals were captured by the SQ 120 system, and cytokines concentrations (pg/mL) were calculated following manufacturer’s instruction using the standard concentrations provided by the kit.

### Cell cultures

#### Human fibroblast cultures

Human fibroblasts were provided by the CCXDP at Massachusetts General Hospital. In this study, we used two fibroblast lines derived from people who lived with XDP and their unaffected family members that were previously described (Ito et al., 2016; Vaine et al., 2017; Aneichyk et al., 2018). All fibroblasts were grown in Dulbecco’s Modified Medium (DMEM) supplemented with 20% FBS, and 1x penicillin/streptomycin/L-glutamine and kept in an incubator at 37°C, 5% CO_2_.

#### SH-SY5Y cells

Human neuroblastoma SH-SY5Y cells (ATCC® CRL-2266) were cultured in DMEM/Nutrient Mixture F-12 (1:1) supplemented with 10% inactivated FBS, 2 mM L-glutamine, 50 μg/mL streptomycin, and 50 IU/mL penicillin. The cells were kept at 5% CO_2_ at 37°C.

#### Oxidative stress detection

ROS levels were measured in both fibroblasts and SH-SY5Y cells by using CellROX Orange reagent (#C10443; Thermo Fisher Scientific, MA), a fluorogenic probe for measuring oxidative stress in live cells, as previously reported (Petrozziello, Bordt et al., 2022). Briefly, following specific treatments, the cells were incubated with 2.5 μM CellROX for 30 min at 37°C in the dark. Then, the medium was replaced with fresh culture media before imaging on a BioTek Cytation 5 imaging reader (BioTek, VT). Images were captured from 6 random areas of each well every 10 min for 2 h. To analyze data generated by Cytation, images were processed using a custom Fiji macro to automatically calculate mean fluorescent intensity (MFI) for each image. The MFI values were then grouped by using a custom Python script, as previously reported (Petrozziello, Bordt et al., 2022). Statistics and XY curves were generated using GraphPad Prism 10.2.0.

#### MPO activity assay

MPO activity was measured by using the OxiSelect Myeloperoxidase Chlorination Activity Assay Kit (#STA-803; Cell Biolabs, CA), according to manufacturer’s protocol. Briefly, samples were incubated in 1 mM hydrogen peroxide (H_2_O_2_) solution for 1 h at RT. Next, samples were incubated in 1x stop solution for 15 min at RT before incubating in 1 mM chromogen working solution for additional 15 min in the dark at RT. All solutions were provided by the kit. At the end of the last incubation, a standard curve was generated following manufacturer’s instruction, and absorbance was read at 405 nm. MPO activity in milliunits/mL (mU/mL) was determined for each sample by dividing the quantity of chromogen consumed by the reaction time as indicated by the kit’s instruction.

#### MPO immuno-depletion

MPO content was immuno-depleted from 4 XDP PFC homogenates that contained the highest MPO levels, as determined by previously published studies (Petrozziello et al., 2020). Specifically, 150 μg of proteins from each XDP PFC was incubated with 15 μL anti-MPO antibody (#A1374, ABclonal Technology, MA) in GAL4 immunoprecipitation buffer, composed of 250 mM sodium chloride (NaCl), 5 mM ethylenediaminetetraacetic acid (EDTA), 1% Nonidet P-40, 50 mM Tris, pH 7.5. After incubation at 4°C for 3.5 h, magnetic protein A beads (Invitrogen, Thermo Fisher, MA) were incubated (20 μL per sample) in agitation overnight at 4°C. At the end of the incubation, all samples were placed on a magnetic rack to separate the supernatants, depleted of MPO. The supernatants were then collected and saved in new tubes labeled as XDP(-). The magnetic beads bound to MPO were, instead, resuspended in GAL4 buffer (50 μL per sample), boiled at 95°C for 5 min and magnetized again to remove the magnetic beads and save the supernatants, enriched in MPO, in new tubes labeled as XDP(+). The efficiency of the immuno-depletion was determined by measuring MPO activity in whole cell extract from XDP, XDP(-) and XDP(+) PFC.

### Cell cultures treatment

#### XDP PFC treatment

SH-SY5Y cells were incubated with 10 ng/mL of homogenates from either XDP PFC (n = 4), XDP(-) (n = 4) or XDP(+) (n = 4) for 24 h before measuring MPO activity and ROS as described above.

#### Verdiperstat

Control– and XDP-derived fibroblasts were treated with either 0.1, 0.5, 1, 5, or 10 μg/mL of verdiperstat for 24 h before measuring MPO activity. ROS levels were measured in control– and XDP-derived fibroblasts in the absence or in the presence of 5 μg/mL of verdiperstat. Similarly, MPO activity and ROS were assessed in XDP-treated SH-SY5Y cells in the absence or in the presence of 5 μg/mL of verdiperstat.

#### Statistics

Normal distribution of data was not assumed regardless of sample size or variance. Individual value plots with the central line representing the median and the whiskers representing the interquartile range, box plot with the central line representing the median, the edges representing the interquartile range, and the whiskers representing the minimum and maximum values, and XY curves were used for graphical representation. Comparisons between groups were performed using a non-parametric Mann-Whitney U test, a one-way ANOVA followed by Tukey’s test, and a two-way ANOVA followed by Tukey’s test. Correlation of MPO levels with XDP clinical features, including age at disease onset, age at death, disease duration and repeat size within the SVA, were performed as non-parametric Spearman correlations. All tests were two-sided with a significance level of 0.05, and exact p values were reported. GraphPad Prism 10.2.0 was used to perform statistical analyses and generate graphs.

#### Study approval

The study was approved by the Partners Healthcare Institutional Review Board at Massachusetts General Hospital (Boston, MA, USA) and by the Makati Medical Center (Makati City, Philippines). Post-mortem consent was obtained from the appropriate representative (next of kin or health care proxy) prior to autopsy.

## Results

### MPO activity was increased in XDP post-mortem PFC

Given the previously reported increase in MPO levels in XDP PFC (Petrozziello et al., 2020), we measured MPO activity in brain tissue sections ex vivo using a specific MPO activity fluorescent imaging probe (MAFA) in a large cohort of post-mortem XDP PFC (Motlagh et al., 2024). Our results revealed a significant increase in MPO activity in XDP PFC compared to controls (Figure 1), consistent with our previously published findings. To determine whether there was an association between known XDP clinical parameters and MPO activity, we correlated MPO activity in PFC with age at disease onset, age at death, disease duration and the size of the hexameric repeat expansion within the SVA in *TAF1* for all XDP samples. The results demonstrated no impact or correlation of any of the clinical parameters with MPO activity in XDP (Figure 2).

**Figure 1.**
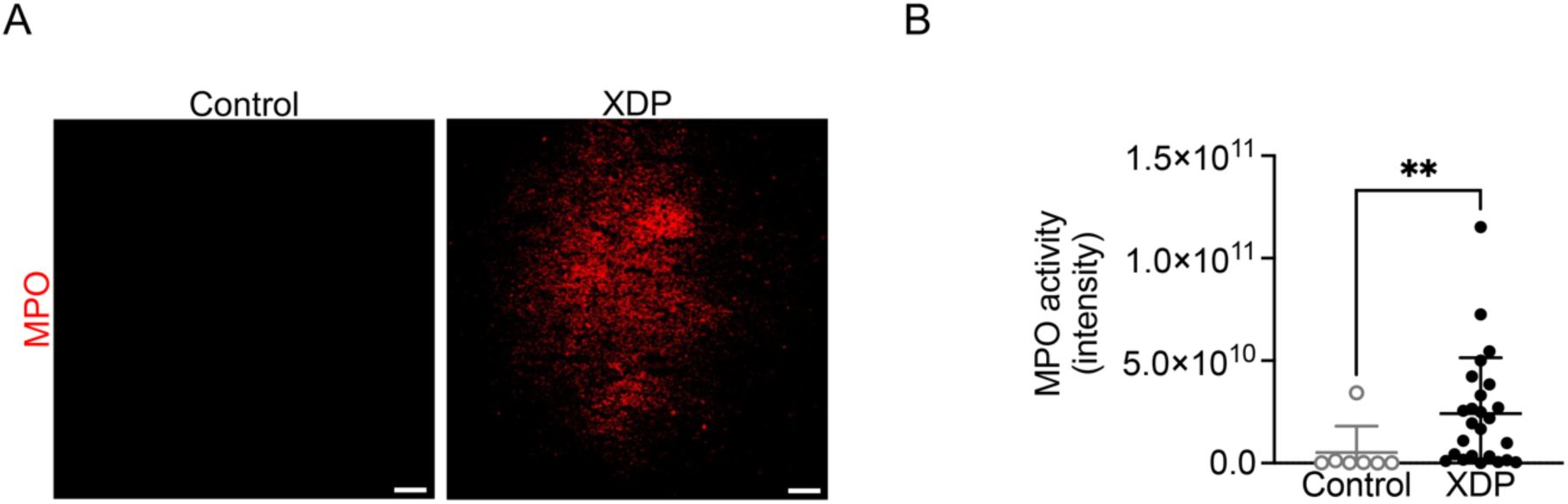
MPO activity was increased in human post-mortem XDP PFC. (**A**) Representative images of MPO activity in control and XDP post-mortem PFC. (**B**) MPO activity was significantly increased in XDP PFC (n = 25) compared to controls (n = 7) (Mann-Whitney U test = 28, p = 0.0051). ** p < 0. 01.

**Figure 2.**
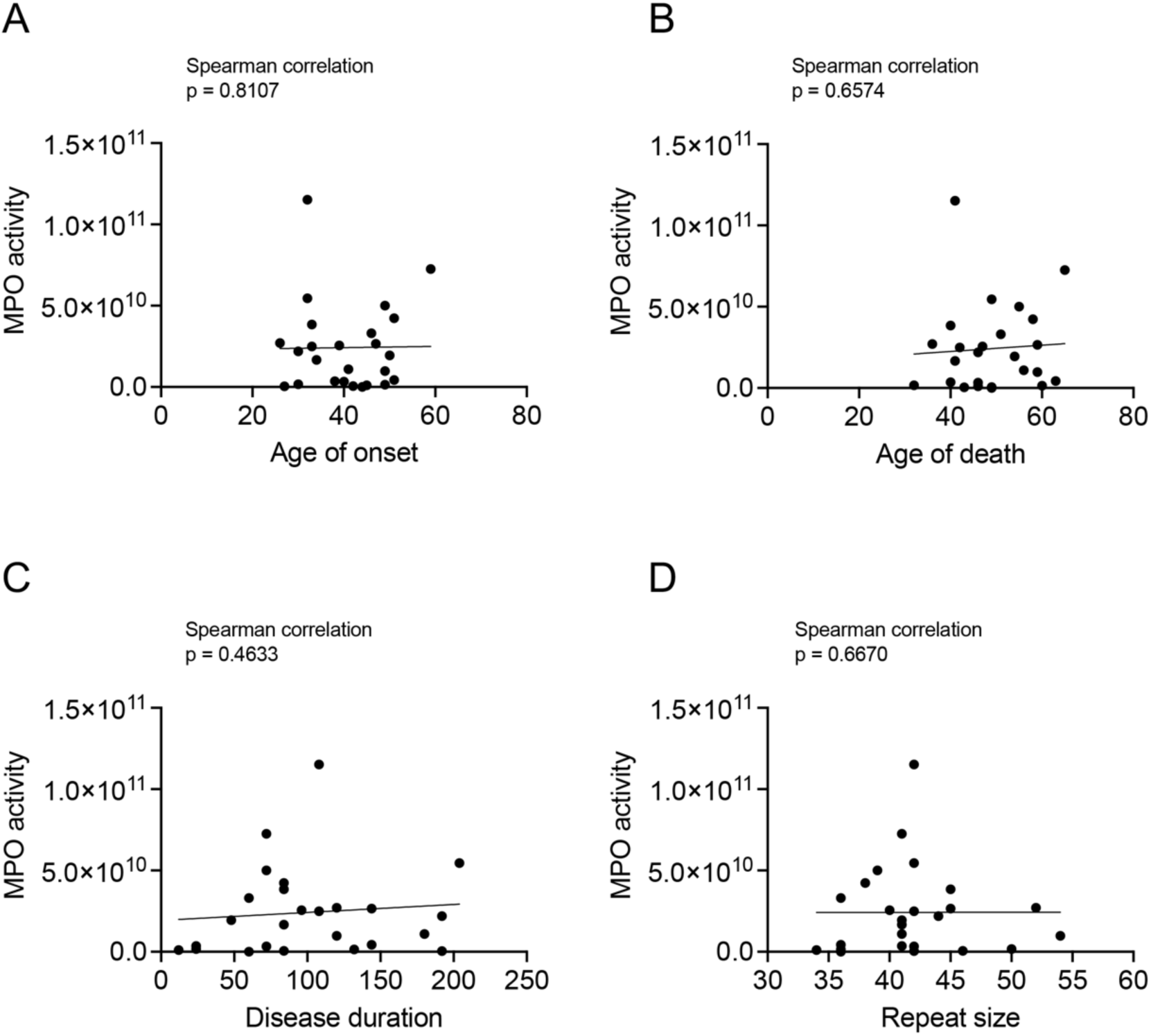
There were no correlations between MPO activity and XDP clinical information. There was no impact of (**A**) age at disease onset (Spearman correlation, r = 0.05046, p = 0.8107), (**B**) age at death (Spearman correlation, r = 0.09327, p = 0.6574), (**C**) disease duration (in months) (Spearman correlation, r = 0.1537, p = 0.4633), or (**D**) repeat size within the SVA (Spearman correlation, r = 0.09051, p = 0.6670) on MPO activity in XDP PFC (n = 25).

### Cytokines levels were not changed in XDP post-mortem PFC

Given that increases in MPO result in increases in pro-inflammatory cytokines (Nussbaum et al., 2013; Khan et al., 2018; Yu et al., 2020), we measured alterations in a panel of 10 cytokines in control and XDP post-mortem PFC using an MSD assay. Our results demonstrated that the levels of INFγ, TNF-α, IL-1β, IL-2, IL-4, IL-6, IL-8, IL-10, IL-12p70, and IL-13 were not significantly different between XDP and control PFC (Supplementary Figure 1).

**Supplementary figure 1.**
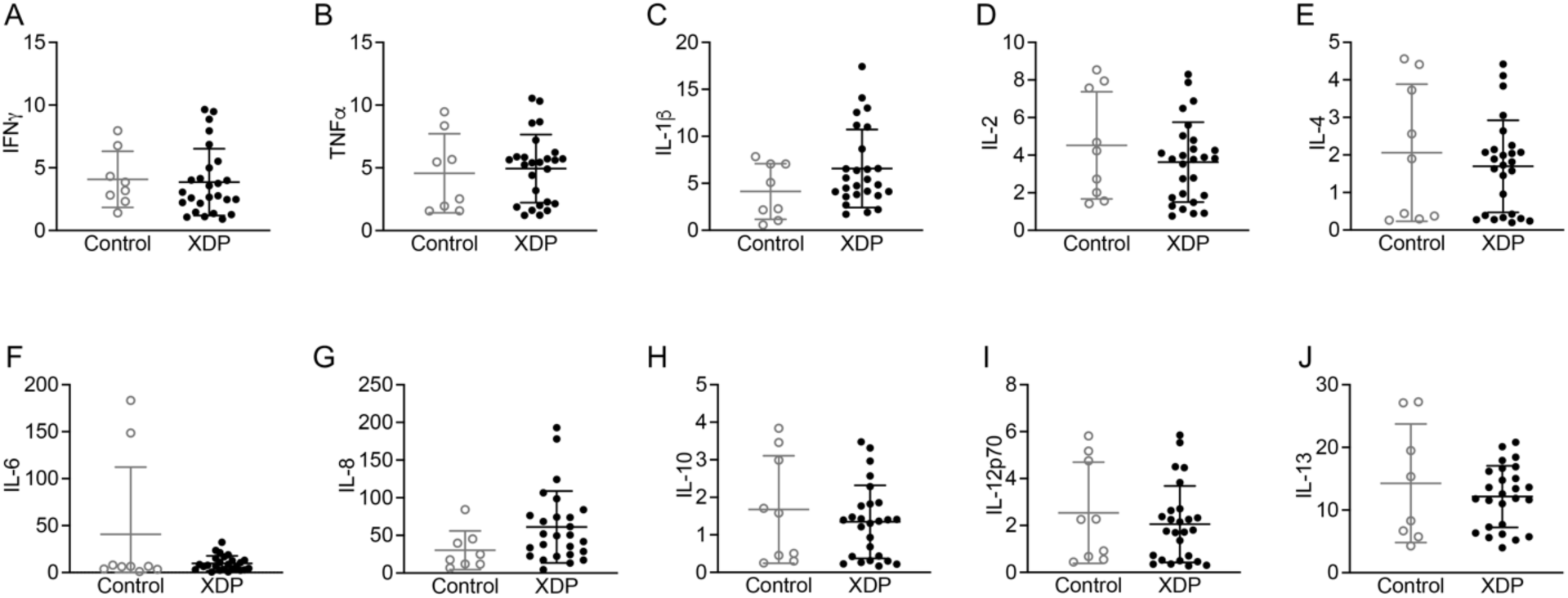
Cytokines levels were not changed in XDP PFC. There were no change in (**A**) IFNγ (Mann-Whitney U test = 89, p = 0.5633), (**B**) TNFα (Mann-Whitney U test = 93, p = 0.6759), (**C**) IL-1β (Mann-Whitney U test = 75, p = 0.2528), (**D**) IL-2 (Mann-Whitney U test = 98, p = 0.4075), (**E**) IL-4 (Mann-Whitney U test = 104, p = 0.6429), (**F**) IL-6 (Mann-Whitney U test = 111, p = 0.9694), (**G**) IL-8 (Mann-Whitney U test = 58, p = 0.0639), (**H**) IL-10 (Mann-Whitney U test = 100, p = 0.5401), (**I**) IL-12p70 (Mann-Whitney U test = 101, p = 0.5650), (**J**) IL-13 (Mann-Whitney U test = 95, p = 0.7351) between XDP (n = 27) and control PFC (n = 9) as measured by MSD assay.

### ROS levels are increased in XDP

Increases in MPO activity are also directly linked to increases in ROS by generating hypochlorous acid (Shaeib et al., 2013; Babior, 2000). Therefore, we measured ROS in XDP-derived fibroblasts by using CellROX and live cell imaging. Antimycin A, an inhibitor of the mitochondrial complex III capable of increasing ROS (Wang et al., 2019), was used as positive control. As expected, ROS levels were significantly increased in antimycin A-treated cells compared to control-derived fibroblasts. Importantly, our results demonstrated a significant increase in ROS in XDP-derived fibroblasts compared to control-derived fibroblasts (Figure 3A).

**Figure 3.**
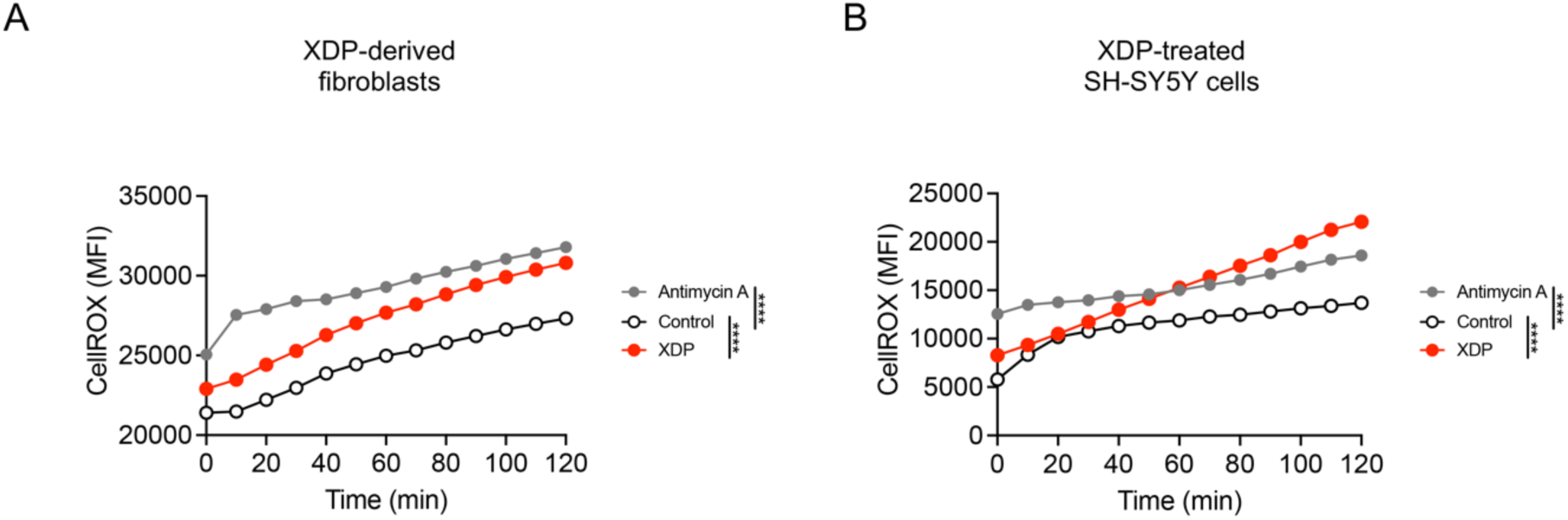
ROS levels were increased in XDP-derived fibroblasts and XDP-treated SH-SY5Y cells. (**A**) Two-way ANOVA demonstrated a significant effect of genotype ([F(2, 24) = 244.5], p < 0.0001), and time ([F(12, 24) = 47.84], p < 0.0001) on ROS levels in XDP fibroblasts. As revealed by Tukey’s test, ROS levels were increased in antimycin A-treated fibroblasts compared to vehicle-treated control fibroblasts (p < 0.0001). Additionally, there was a significant increase in ROS in vehicle-treated XDP fibroblasts compared to control fibroblasts (p < 0.0001). (**B**) Two-way ANOVA demonstrated a significant effect of treatment ([F(2, 24) = 25.66], p < 0.0001), and time ([F(12, 24) = 8.994], p < 0.0001) on ROS levels in SH-SY5Y cells. As expected, Tukey’s test revealed a significant increase in ROS in antimycin A-treated cells compared to vehicle-treated cells (p < 0.0001). Importantly, there was a significant increase in ROS in XDP-treated cells compared to vehicle-treated cells (p < 0.0001). **** p < 0.0001

Next, we sought to determine whether the treatment of SH-SY5Y cells with whole cell extracts derived from XDP PFC could recapitulate pathological features of XDP, such as increases in oxidative stress, as we have recently demonstrated in ALS (Petrozziello, Bordt et al., 2022). SH-SY5Y cells were treated with XDP PFC (10 ng/mL) for 24 h before ROS levels were measured as described above. As expected, antimycin A induced a significant increase in ROS compared to vehicle-treated cells. Furthermore, there was a significant increase in ROS in XDP-treated SH-SY5Y cells compared to vehicle-treated cells (Figure 3B).

### MPO immuno-depletion reduced MPO activity and ROS in vitro

To determine whether increases in MPO directly induce an increase in ROS in XDP, we removed MPO from XDP post-mortem PFC homogenates by performing an immuno-depletion experiment. To determine the efficiency of immuno-depletion, MPO activity was measured in whole cell extracts from XDP PFC, XDP PFC depleted of MPO [XDP(-)], and XDP PFC enriched in MPO [XDP(+)] by using a commercially available kit. The results revealed that MPO activity was significantly decreased in XDP(-) compared to both XDP(+) and XDP PFC, confirming the successful removal of MPO from the samples (Figure 4A).

**Figure 4.**
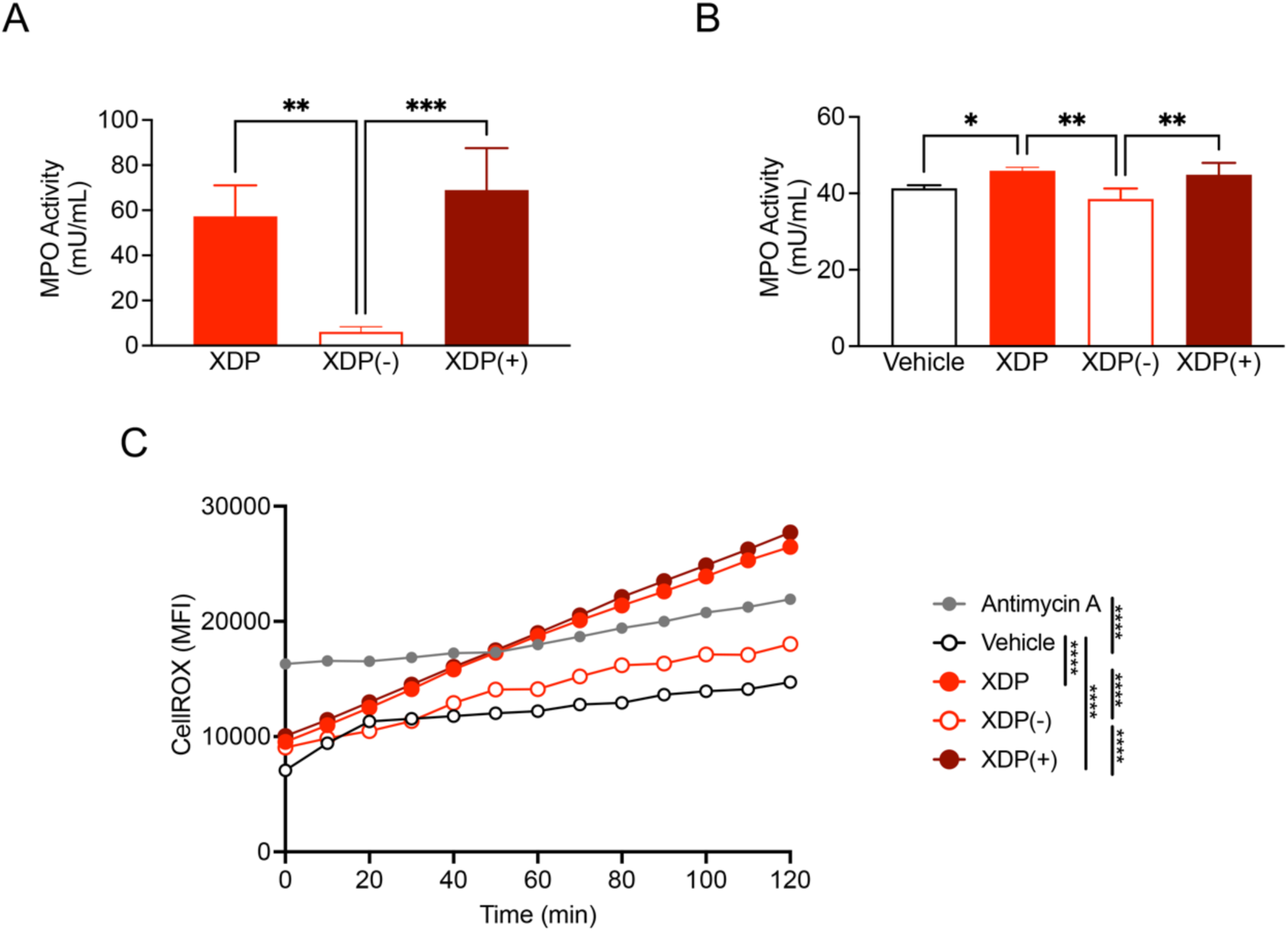
MPO immuno-depletion decreased ROS levels in SH-SY5Y cells. (**A**) One-way ANOVA revealed a significant effect of immuno-depletion ([F(2, 9) = 24.7], p = 0.0002) on MPO activity in SH-SY5Y cells. Specifically, Tukey’s test revealed a significant decrease in MPO activity in XDP(-) treated cells compared to both XDP (p = 0.011), and XDP(+) treated cells (p = 0.0003). (**B**) One-way ANOVA demonstrated a significant effect of treatment ([F(3, 12) = 9.763], p = 0.0015) on MPO activity in SH-SY5Y cells. Tukey’s test demonstrated a significant increase in MPO activity in XDP treated cells compared to vehicle treated cells (p = 0.0460) as well as a significant decrease in MPO activity in XDP(-) cells compared to both XDP (p = 0.0020) and XDP(+) cells (p = 0.0066). (**C**) Two-way ANOVA revealed a significant effect of immuno-depletion ([F(4, 48) = 32.85], p < 0.0001) and time ([F(12, 48) = 16.96], p < 0.0001) on ROS levels in SH-SY5Y cells. As expected, there was an increase in ROS in antimycin A treated cells compared to vehicle treated cells (Tukey’s test, p < 0.0001). Additionally, Tukey’s test demonstrated a significant increase in ROS in XDP (p < 0.0001) and XDP(+) treated cells (p < 0.0001) compared to vehicle treated cells. Importantly, Tukey’s test also revealed a significant decrease in ROS in XDP(-) treated cells compared to both XDP (p < 0.0001) and XDP(+) cells (p < 0.0001). * p < 0.05; ** p < 0.01; *** p < 0.001; **** p < 0.0001.

Next, SH-SY5Y cells were treated with XDP, XDP(-), and XDP(+) PFC (10 ng/mL) for 24 h before measuring MPO activity using a commercially available kit. Our findings indicated a significant increase in MPO activity in XDP and XDP(+) treated cells compared to vehicle treated cells. Importantly, MPO activity was significantly decreased in XDP(-) treated cells (Figure 4B).

Lastly, to determine whether MPO causes increase in ROS in XDP, ROS levels were measured in SH-SY5Y cells following treatment with either XDP, XDP(-), or XDP(+) PFC (10 ng/mL/24h) by using CellROX and live cell imaging. As expected, the positive control antimycin A induced a significant increase in ROS. A similar increase was measured in XDP and XDP(+) treated cells compared to vehicle treated cells. Importantly, there was a significant decrease in ROS in XDP(-) treated cells compared to both XDP and XDP(+) treated cells (Figure 4C).

### Verdiperstat reduced MPO activity and ROS in XDP-derived fibroblasts

Next, we assessed the effects of a potent and selective MPO inhibitor, verdiperstat, on MPO activity in control– and XDP-derived fibroblasts. The cells were treated with verdiperstat (0.1, 0.5, 1, 5, and 10 μg/mL), for 24 h and MPO activity was measured with a commercially available kit. Verdiperstat treatment significantly decreased MPO activity in a dose-dependent manner. Specifically, there was a significant decrease in MPO activity in XDP-derived fibroblasts treated with either 1, 5 or 10 μg/mL of verdiperstat compared to vehicle-treated XDP-derived fibroblasts (Figure 5A). Based on these results, we used 5 μg/mL of verdiperstat for the next set of experiments outlined below given that this dose decreased MPO activity by 50%.

**Figure 5.**
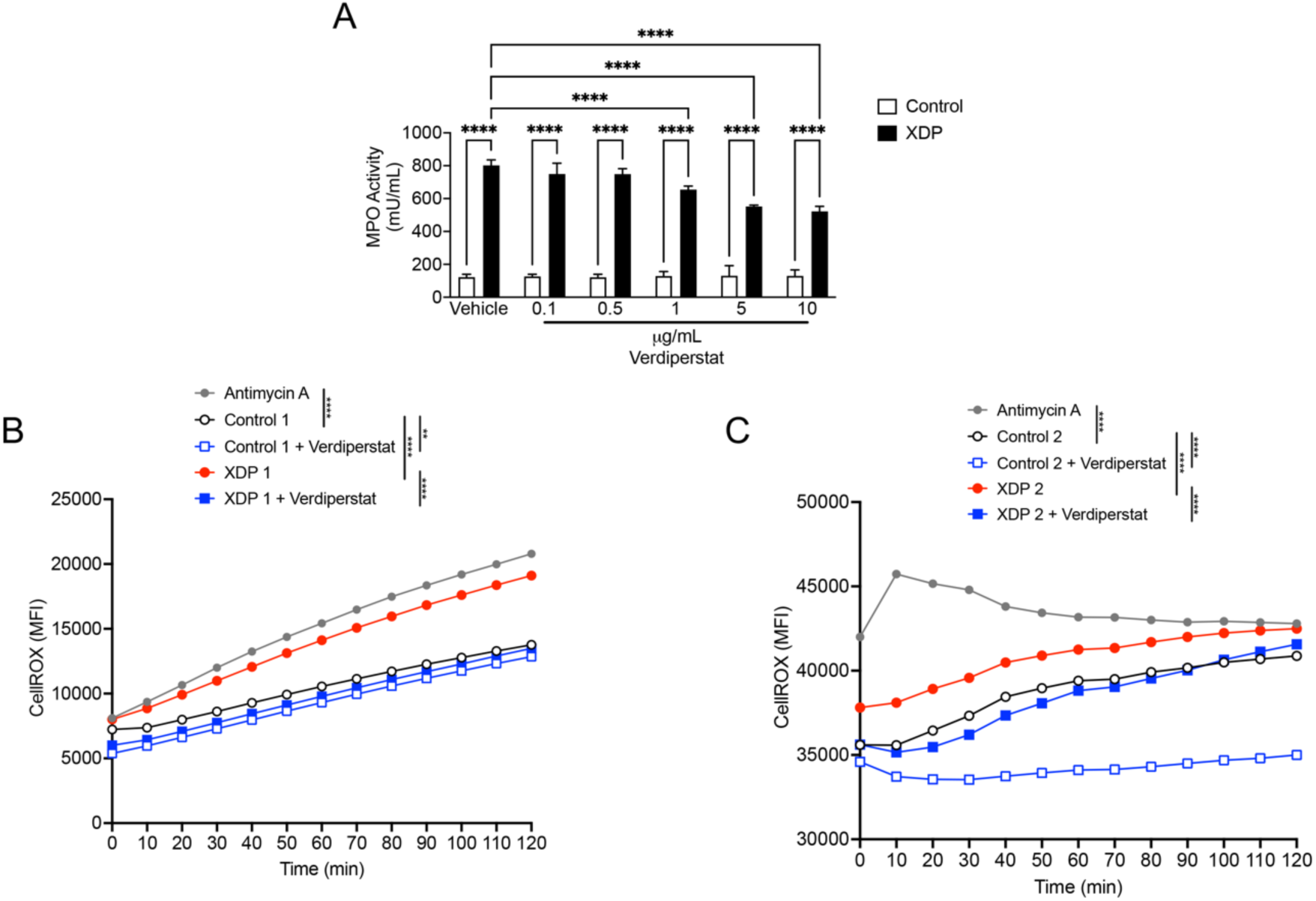
Verdiperstat decreased MPO activity and ROS levels in XDP fibroblasts. (**A**) Two-way ANOVA revealed a significant effect of treatment ([F(5, 36) = 20.08], p < 0.0001), genotype ([F(1, 36) = 2890], p<0.0001), and treatment x XDP interaction ([F(5, 36) = 22.71], p < 0.0001) on MPO activity in fibroblasts. Tukey’s’ test demonstrated a significant increase in MPO activity in XDP-compared to control-derived fibroblasts (p < 0.0001) as well as a significant increase in MPO activity in XDP-derived fibroblasts treated with either 0.1 (p < 0.0001), 0.5 (p < 0.0001), 1 (p < 0.0001), 5 (p < 0.0001), or 10 μg/mL of verdiperstat (p < 0.0001) compared to control-derived fibroblasts treated with the same doses of verdiperstat. Lastly, Tukey’s test revealed a significant decrease in MPO activity in XDP-derived fibroblasts treated with either 5 (p < 0.0001) or 10 μg/mL of verdiperstat (p < 0.0001) compared to vehicle-treated XDP-derived fibroblasts. **(B)** Two-way ANOVA revealed a significant effect of genotype ([F(4, 48) = 120.3], p < 0.001), and time ([F(12, 48) = 61.32], p < 0.0001) on ROS levels in fibroblasts. Tukey’s test revealed an expected significant increase in ROS in antimycin A treated fibroblasts compared to vehicle-treated control fibroblasts (p < 0.0001). There was also a significant increase in ROS in XDP fibroblasts compared to control fibroblasts (p < 0.0001). Importantly, Tukey’s test demonstrated a significant decrease in ROS in both control– and XDP-derived fibroblasts treated with verdiperstat compared to vehicle-treated control (p = 0.052) and XDP fibroblasts (p < 0.0001). **(C)** Two-way ANOVA demonstrated a significant effect of genotype ([F(4, 48) = 91.45], p < 0.0001), and time ([F(12, 48) = 3.579], p = 0.0008) on ROS levels in a second set of fibroblasts. As expected, there was a significant increase in antimycin A treated-fibroblasts compared to vehicle-treated control fibroblasts (p < 0.0001). Furthermore, Tukey’s test demonstrated a significant increase in ROS in XDP fibroblasts compared to control fibroblasts (p = 0.0025). Importantly, Tukey’s test revealed a significant decrease in ROS levels in both control– and XDP-derived fibroblasts treated with verdiperstat compared to vehicle-treated control (p < 0.0001) and XDP fibroblasts (p = 0.0002). ** p < 0.001; **** p<0.0001.

Next, the effect of verdiperstat on ROS levels was measured in control– and XDP-derived fibroblasts using CellROX and live cell imaging as described above. The results confirmed a significant increase in ROS in XDP-derived fibroblasts compared to control-derived fibroblasts (Figure 5B and C). Importantly, verdiperstat treatment reduced ROS in both control– and XDP-derived fibroblasts (Figure 5B and C).

### Verdiperstat reduced MPO activity and ROS in XDP-treated SH-SY5Y cells

To verify whether verdiperstat was also able to reduce MPO activity and ROS in SH-SY5Y cells treated with post-mortem XDP PFC, the cells were treated with XDP PFC homogenates (10 ng/mL) for 24 h in the absence or in the presence of verdipestat (5 μg/mL) and MPO activity was measured as previously described. The results confirmed a significant increase in MPO activity in XDP-treated SH-SY5Y cells compared to vehicle-treated cells. Importantly, there was a significant decrease in MPO activity in XDP+verdiperstat treated SH-SY5Y cells compared to XDP-treated SH-SY5Y cells (Figure 6A).

**Figure 6.**
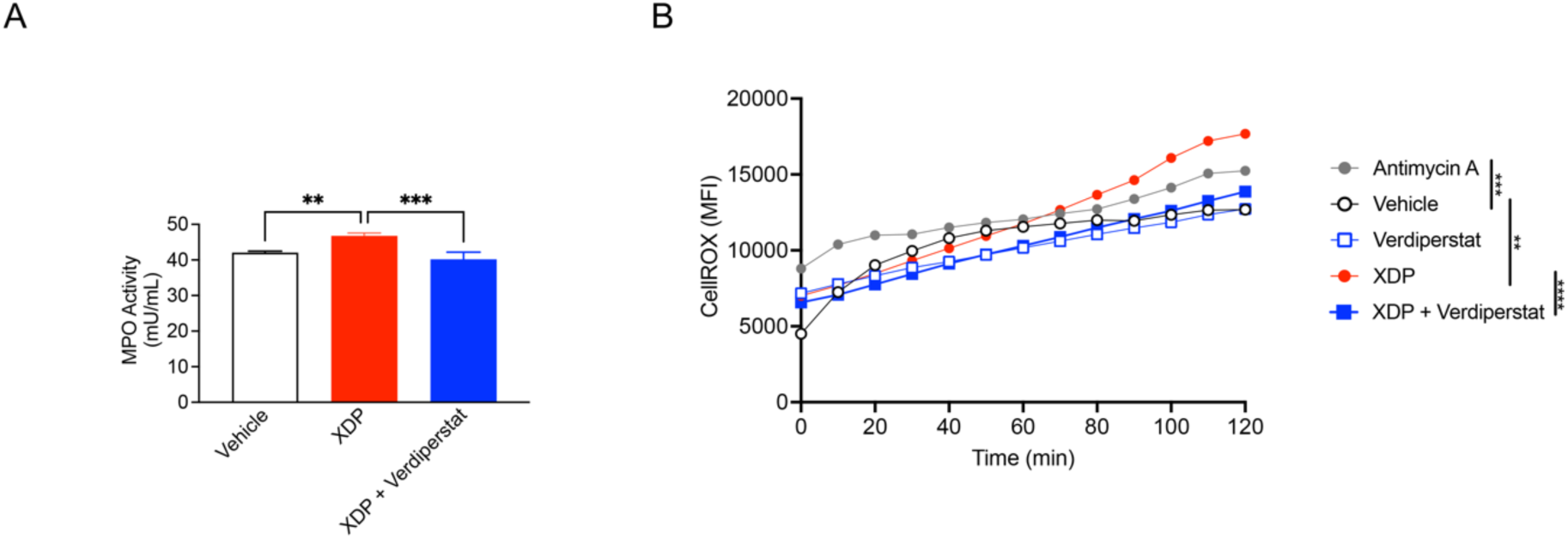
Verdiperstat decreased MPO activity and ROS levels in XDP-treated SH-SY5Y cells. (**A**) One-way ANOVA revealed a significant effect of treatment ([F(2, 9) = 29.19], p = 0.0001) on MPO activity in SH-SY5Y cells. Tukey’s test revealed a significant increase in MPO activity in XDP treated cells compared to both vehicle (p = 0.0012) and XDP+verdiperstat treated cells (p=0.0001). **(B)** Two-way ANOVA revealed a significant effect of treatment ([F(4, 48) = 16.35], p < 0.0001), and time ([F(12, 48) = 31.68], p < 0.001) on ROS levels in SH-SY5Y cells. As expected, ROS levels were increased in antimycin A treated cells compared to vehicle treatment (Tukey’s test, p = 0.0003). Additionally, Tukey’s test demonstrated a significant increase in ROS in XDP treated cells compared to both vehicle (p = 0.0014) and XDP+verdiperstat treated cells (p < 0.0001, respectively). ** p<0.01; *** p<0.001; **** p<0.0001.

Lastly, we measured ROS in SH-SY5Y cells following treatment with XDP (10 ng/mL) for 24 h in the absence or in the presence of verdiperstat (5μg/mL) using CellROX and live cell imaging. The results demonstrated a significant increase in ROS in SH-SY5Y cells treated with the positive control antimycin A as well as in XDP treated cells compared to vehicle treated cells. Importantly, there was a significant decrease in ROS in XDP+verdiperstat treated SH-SY5Y cells compared to XDP treated cells (Figure 6B).

## Discussion

In this study, we demonstrated a significant increase in MPO activity ex vivo in a large cohort of XDP human post-mortem PFC using a novel MPO fluorescent imaging probe (Motlagh et al., 2024). While our results revealed that increases in MPO did not alter pro– and anti-inflammatory cytokine levels, it contributed to a significant increase in ROS in XDP. Both immuno-depletion of MPO as well as inhibition of MPO using verdiperstat reduced MPO activity and resulted in significant decreases in ROS in XDP-derived fibroblasts and SH-SY5Y cells treated with post-mortem XDP PFC. Collectively, these findings suggest that inhibiting MPO dampens oxidative stress in XDP.

Recently, we demonstrated a significant increase in neuroinflammation and MPO in XDP post-mortem PFC (Petrozziello et al., 2020), highlighting MPO as a contributor to this neuroinflammatory process similar to other neurodegenerative diseases. Indeed, increases in MPO levels, astrogliosis and microgliosis have been described in several neurodegenerative diseases, including AD (Reynolds et al., 1999; Green et al., 2004; Gelhaar et al., 2017), PD (Choi et al., 2005; Jucaite et al., 2015; Gelhaar et al., 2017) and multiple sclerosis (MS) (Nagra et al., 1997; Minohara et al., 2006; Gray et al., 2008).

MPO plays a critical role in the innate immune response as it contributes to both neutrophil antimicrobial activity and phagocytosis (Nicholls et al., 2005); however, a prolonged increase in the systemic levels of MPO could cause extravasation into extracellular spaces, causing tissue damage during chronic inflammation (Parker et al., 2012; Hawkins and Davies, 2021). One consequence of the sustained increase in MPO is the rise in the expression and release of pro-inflammatory cytokines (Nussbaum et al., 2013; Khan et al., 2018; Yu et al., 2020). However, our findings suggest that increases in MPO levels and activity in XDP, specifically, did not alter cytokine levels. It should be noted that one caveat of the current study is that we only assessed a panel of 10 cytokines and that we did not fully characterize the effects of MPO on a complete panel of inflammatory markers. Therefore, it is possible that increases in MPO may alter the expression and release of other cytokines or chemokines such as CXCL13 and CX3CL1 as we have previously demonstrated in XDP-derived fibroblasts (Petrozziello et al., 2020). Another downstream consequence of increased MPO is oxidative stress and an increase in ROS production through the generation of hypochlorous acid (Shaeib et al., 2013; Babior, 2000). Here, we report a significant increase in ROS in both XDP-derived fibroblasts and XDP PFC-treated SH-SY5Y cells. Furthermore, the increase in ROS is a direct consequence of increased MPO levels in XDP as it was mitigated by decreasing MPO by either immuno-depletion or treatment with verdiperstat. Together these findings demonstrate that inhibiting MPO could represent a viable therapeutic strategy to dampen oxidative stress in XDP, although future studies need to determine if decreasing ROS could prevent or reduce neurodegeneration and demonstrate clinical benefit.

Verdiperstat is a first-in-class, potent, selective, brain-permeable MPO inhibitor produced by Biohaven Pharmaceuticals, Inc. Treatment with verdiperstat has been shown to decrease microglia activation in multiple system atrophy (MSA) (Stefanova et al., 2012) and PD (Jucaite et al., 2015). In addition, verdiperstat has been tested in two clinical trials demonstrating safety and tolerability in people (NCT03952806 or M-STAR study in multiple system atrophy, and NCT04436510 or Regimen B in the Healey ALS Platform Trial in ALS). Although these trials failed to meet the primary endpoints, the potential beneficial effect of verdiperstat in treating neurodegenerative diseases is still under investigation and a third clinical trial (Veri-T) is currently ongoing to determine the therapeutic efficacy of verdiperstat for the treatment of semantic variant primary progressive aphasia (svPPA) (NCT05184569). Therefore, our findings in XDP may pave the way for further assessing the potential beneficial therapeutic effect of verdiperstat in dampening neurodegeneration in XDP.

## Conclusion

In summary, our findings demonstrate that increases in MPO may contribute to XDP pathogenesis, as revealed by a significant increase in MPO activity ex vivo in human post-mortem XDP PFC. Additionally, our results indicate that MPO causes oxidative stress in XDP, as demonstrated by increases in ROS in cellular models of XDP. Importantly, depleting MPO or using a selective and potent MPO inhibitor mitigated ROS levels in XDP and lays the foundation for future studies to assess the neuroprotective effects of MPO inhibition in XDP.

## Author contributions

T.P. contributed to conceptualization, data curation, formal analysis, funding acquisition, investigation, methodology, software, supervision, validation, visualization, writing – original draft; N.J.M., R.Z.B.M., D.L., and M.G.M. contributed to data curation, formal analysis, visualization, and writing – review and editing. E.B.P., D.C.B., C.F.-C., G.P.L., M.S., E.M., M.C.A., C.C.E.D., C.Z., R.E.T., I.A.Q., and J.W.C. contributed to supervision and writing – review and editing.; G.S.-V. contributed conceptualization, data curation, funding acquisition, investigation, project administration, resources, supervision, validation, visualization, writing – original draft.

## Acknowledgments

The authors would like to thank patients and their family for sample donation.

## Data availability statement

The data supporting the findings of this study are available within the article and/or its supplementary material.

## References

1. Al Ali, J., Vaine, C. A., Shah, S., Campion, L., Hakoum, A., Supnet, M. L., Acuna, P., Aldykiewicz, G., Multhaupt-Buell, T., Ganza, N. G. M., Lagarde, J. B. B., De Guzman, J. K., Go, C., Currall, B., Trombetta, B., Webb, P. K., Talkowski, M., Arnold, S. E., Cheah, P. S., … Breakefield, X. O. (2021). TAF1 Transcripts and Neurofilament Light Chain as Biomarkers for X-linked Dystonia-Parkinsonism. Mov Disord, 36(1), 206–215. 10.1002/mds.28305

2. Aneichyk, T., Hendriks, W. T., Yadav, R., Shin, D., Gao, D., Vaine, C. A., Collins, R. L., Domingo, A., Currall, B., Stortchevoi, A., Multhaupt-Buell, T., Penney, E. B., Cruz, L., Dhakal, J., Brand, H., Hanscom, C., Antolik, C., Dy, M., Ragavendran, A., … Talkowski, M. E. (2018). Dissecting the Causal Mechanism of X-Linked Dystonia-Parkinsonism by Integrating Genome and Transcriptome Assembly. Cell, 172(5), 897–909 e821. 10.1016/j.cell.2018.02.011

3. Araujo, B., Caridade-Silva, R., Soares-Guedes, C., Martins-Macedo, J., Gomes, E. D., Monteiro, S., & Teixeira, F. G. (2022). Neuroinflammation and Parkinson’s Disease-From Neurodegeneration to Therapeutic Opportunities. Cells, 11(18). 10.3390/cells11182908

4. Babior, B. M. (2000). Phagocytes and oxidative stress. Am J Med, 109(1), 33–44. 10.1016/s0002-9343(00)00481-2

5. Bragg, D. C., Mangkalaphiban, K., Vaine, C. A., Kulkarni, N. J., Shin, D., Yadav, R., Dhakal, J., Ton, M. L., Cheng, A., Russo, C. T., Ang, M., Acuna, P., Go, C., Franceour, T. N., Multhaupt-Buell, T., Ito, N., Muller, U., Hendriks, W. T., Breakefield, X. O., … Ozelius, L. J. (2017). Disease onset in X-linked dystonia-parkinsonism correlates with expansion of a hexameric repeat within an SVA retrotransposon in TAF1. Proc Natl Acad Sci U S A, 114(51), E11020–E11028. 10.1073/pnas.1712526114

6. Choi, D. K., Pennathur, S., Perier, C., Tieu, K., Teismann, P., Wu, D. C., Jackson-Lewis, V., Vila, M., Vonsattel, J. P., Heinecke, J. W., & Przedborski, S. (2005). Ablation of the inflammatory enzyme myeloperoxidase mitigates features of Parkinson’s disease in mice. J Neurosci, 25(28), 6594–6600. 10.1523/JNEUROSCI.0970-05.2005

7. Domingo, A., Westenberger, A., Lee, L. V., Braenne, I., Liu, T., Vater, I., Rosales, R., Jamora, R. D., Pasco, P. M., Cutiongco-Dela Paz, E. M., Freimann, K., Schmidt, T. G., Dressler, D., Kaiser, F. J., Bertram, L., Erdmann, J., Lohmann, K., & Klein, C. (2015). New insights into the genetics of X-linked dystonia-parkinsonism (XDP, DYT3). Eur J Hum Genet, 23(10), 1334–1340. 10.1038/ejhg.2014.292

8. Fernandez-Cerado, C., Legarda, G. P., Velasco-Andrada, M. S., Aguil, A., Ganza Bautista, N. G., Lagarde, J. B. B., Soria, J., Jamora, R. D. G., Acuna, P. J., Vanderburg, C., Sapp, E., DiFiglia, M., Murcar, M. G., Campion, L., Ozelius, L. J., Alessi, A. K., Singh-Bains, M. K., Waldvogel, H. J., Faull, R. L. M., … Acuna-Sunshine, G. (2021). Promise and challenges of dystonia brain banking: establishing a human tissue repository for studies of X-Linked Dystonia-Parkinsonism. J Neural Transm (Vienna), 128(4), 575–587. 10.1007/s00702-020-02286-9

9. Forghani, R., Kim, H. J., Wojtkiewicz, G. R., Bure, L., Wu, Y., Hayase, M., Wei, Y., Zheng, Y., Moskowitz, M. A., & Chen, J. W. (2015). Myeloperoxidase propagates damage and is a potential therapeutic target for subacute stroke. J Cereb Blood Flow Metab, 35(3), 485–493. 10.1038/jcbfm.2014.222

10. Gellhaar, S., Sunnemark, D., Eriksson, H., Olson, L., & Galter, D. (2017). Myeloperoxidase-immunoreactive cells are significantly increased in brain areas affected by neurodegeneration in Parkinson’s and Alzheimer’s disease. Cell Tissue Res, 369(3), 445–454. 10.1007/s00441-017-2626-8

11. Gray, E., Thomas, T. L., Betmouni, S., Scolding, N., & Love, S. (2008). Elevated activity and microglial expression of myeloperoxidase in demyelinated cerebral cortex in multiple sclerosis. Brain Pathol, 18(1), 86–95. 10.1111/j.1750-3639.2007.00110.x

12. Green, P. S., Mendez, A. J., Jacob, J. S., Crowley, J. R., Growdon, W., Hyman, B. T., & Heinecke, J. W. (2004). Neuronal expression of myeloperoxidase is increased in Alzheimer’s disease. J Neurochem, 90(3), 724–733. 10.1111/j.1471-4159.2004.02527.x

13. Hawkins, C. L., & Davies, M. J. (2021). Role of myeloperoxidase and oxidant formation in the extracellular environment in inflammation-induced tissue damage. Free Radic Biol Med, 172, 633–651. 10.1016/j.freeradbiomed.2021.07.007

14. Ito, N., Hendriks, W. T., Dhakal, J., Vaine, C. A., Liu, C., Shin, D., Shin, K., Wakabayashi-Ito, N., Dy, M., Multhaupt-Buell, T., Sharma, N., Breakefield, X. O., & Bragg, D. C. (2016). Decreased N-TAF1 expression in X-linked dystonia-parkinsonism patient-specific neural stem cells. Dis Model Mech, 9(4), 451–462. 10.1242/dmm.022590

15. Jucaite, A., Svenningsson, P., Rinne, J. O., Cselenyi, Z., Varnas, K., Johnstrom, P., Amini, N., Kirjavainen, A., Helin, S., Minkwitz, M., Kugler, A. R., Posener, J. A., Budd, S., Halldin, C., Varrone, A., & Farde, L. (2015). Effect of the myeloperoxidase inhibitor AZD3241 on microglia: a PET study in Parkinson’s disease. Brain, 138(Pt 9), 2687–2700. 10.1093/brain/awv184

16. Khan, A. A., Alsahli, M. A., & Rahmani, A. H. (2018). Myeloperoxidase as an Active Disease Biomarker: Recent Biochemical and Pathological Perspectives. Med Sci (Basel), 6(2). 10.3390/medsci6020033

17. Lee, L. V., Rivera, C., Teleg, R. A., Dantes, M. B., Pasco, P. M., Jamora, R. D., Arancillo, J., Villareal-Jordan, R. F., Rosales, R. L., Demaisip, C., Maranon, E., Peralta, O., Borres, R., Tolentino, C., Monding, M. J., & Sarcia, S. (2011). The unique phenomenology of sex-linked dystonia parkinsonism (XDP, DYT3, “Lubag”). Int J Neurosci, 121 *Suppl 1*, 3–11. 10.3109/00207454.2010.526728

18. Makino, S., Kaji, R., Ando, S., Tomizawa, M., Yasuno, K., Goto, S., Matsumoto, S., Tabuena, M. D., Maranon, E., Dantes, M., Lee, L. V., Ogasawara, K., Tooyama, I., Akatsu, H., Nishimura, M., & Tamiya, G. (2007). Reduced neuron-specific expression of the TAF1 gene is associated with X-linked dystonia-parkinsonism. Am J Hum Genet, 80(3), 393–406. 10.1086/512129

19. Minohara, M., Matsuoka, T., Li, W., Osoegawa, M., Ishizu, T., Ohyagi, Y., & Kira, J. (2006). Upregulation of myeloperoxidase in patients with opticospinal multiple sclerosis: positive correlation with disease severity. J Neuroimmunol, 178(1-2), 156–160. 10.1016/j.jneuroim.2006.05.026

20. Motlagh, N.J., Wang, C., Kim, H.-H., Jun, Y., Kim, D., Lee, S., Chen, J.W. (2024). Aging intensifies myeloperoxidase activity after ischemic stroke. Aging and Disease *–* in press

21. Nagra, R. M., Becher, B., Tourtellotte, W. W., Antel, J. P., Gold, D., Paladino, T., Smith, R. A., Nelson, J. R., & Reynolds, W. F. (1997). Immunohistochemical and genetic evidence of myeloperoxidase involvement in multiple sclerosis. J Neuroimmunol, 78(1-2), 97–107. 10.1016/s0165-5728(97)00089-1

22. Nicholls, S. J., Zheng, L., & Hazen, S. L. (2005). Formation of dysfunctional high-density lipoprotein by myeloperoxidase. Trends Cardiovasc Med, 15(6), 212–219. 10.1016/j.tcm.2005.06.004

23. Nolte, D., Niemann, S., & Muller, U. (2003). Specific sequence changes in multiple transcript system DYT3 are associated with X-linked dystonia parkinsonism. Proc Natl Acad Sci U S A, 100(18), 10347–10352. 10.1073/pnas.1831949100

24. Nussbaum, J. C., Van Dyken, S. J., von Moltke, J., Cheng, L. E., Mohapatra, A., Molofsky, A. B., Thornton, E. E., Krummel, M. F., Chawla, A., Liang, H. E., & Locksley, R. M. (2013). Type 2 innate lymphoid cells control eosinophil homeostasis. Nature, 502(7470), 245–248. 10.1038/nature12526

25. Obrador, E., Salvador, R., Lopez-Blanch, R., Jihad-Jebbar, A., Valles, S. L., & Estrela, J. M. (2020). Oxidative Stress, Neuroinflammation and Mitochondria in the Pathophysiology of Amyotrophic Lateral Sclerosis. Antioxidants (Basel), 9(9). 10.3390/antiox9090901

26. Parker, H., & Winterbourn, C. C. (2012). Reactive oxidants and myeloperoxidase and their involvement in neutrophil extracellular traps. Front Immunol, 3, 424. 10.3389/fimmu.2012.00424

27. Petrozziello, T., Bordt, E. A., Mills, A. N., Kim, S. E., Sapp, E., Devlin, B. A., Obeng-Marnu, A. A., Farhan, S. M. K., Amaral, A. C., Dujardin, S., Dooley, P. M., Henstridge, C., Oakley, D. H., Neueder, A., Hyman, B. T., Spires-Jones, T. L., Bilbo, S. D., Vakili, K., Cudkowicz, M. E., … Sadri-Vakili, G. (2022). Targeting Tau Mitigates Mitochondrial Fragmentation and Oxidative Stress in Amyotrophic Lateral Sclerosis. Mol Neurobiol, 59(1), 683–702. 10.1007/s12035-021-02557-w

28. Petrozziello, T., Mills, A. N., Vaine, C. A., Penney, E. B., Fernandez-Cerado, C., Legarda, G. P. A., Velasco-Andrada, M. S., Acuna, P. J., Ang, M. A., Munoz, E. L., Diesta, C. C. E., Macalintal-Canlas, R., Acuna-Sunshine, G., Ozelius, L. J., Sharma, N., Bragg, D. C., & Sadri-Vakili, G. (2020). Neuroinflammation and histone H3 citrullination are increased in X-linked Dystonia Parkinsonism post-mortem prefrontal cortex. Neurobiol Dis, 144, 105032. 10.1016/j.nbd.2020.105032

29. Raven, F., Ward, J. F., Zoltowska, K. M., Wan, Y., Bylykbashi, E., Miller, S. J., Shen, X., Choi, S. H., Rynearson, K. D., Berezovska, O., Wagner, S. L., Tanzi, R. E., & Zhang, C. (2017). Soluble Gamma-secretase Modulators Attenuate Alzheimer’s beta-amyloid Pathology and Induce Conformational Changes in Presenilin 1. EBioMedicine, 24, 93–101. 10.1016/j.ebiom.2017.08.028

30. Reynolds, W. F., Rhees, J., Maciejewski, D., Paladino, T., Sieburg, H., Maki, R. A., & Masliah, E. (1999). Myeloperoxidase polymorphism is associated with gender specific risk for Alzheimer’s disease. Exp Neurol, 155(1), 31–41. 10.1006/exnr.1998.6977

31. Saba, J., Couselo, F. L., Bruno, J., Carniglia, L., Durand, D., Lasaga, M., & Caruso, C. (2022). Neuroinflammation in Huntington’s Disease: A Starring Role for Astrocyte and Microglia. Curr Neuropharmacol, 20(6), 1116–1143. 10.2174/1570159X19666211201094608

32. Shaeib, F., Khan, S. N., Thakur, M., Kohan-Ghadr, H. R., Drewlo, S., Saed, G. M., Pennathur, S., & Abu-Soud, H. M. (2016). The Impact of Myeloperoxidase and Activated Macrophages on Metaphase II Mouse Oocyte Quality. PLoS One, 11(3), e0151160. 10.1371/journal.pone.0151160

33. Stefanova, N., Georgievska, B., Eriksson, H., Poewe, W., & Wenning, G. K. (2012). Myeloperoxidase inhibition ameliorates multiple system atrophy-like degeneration in a transgenic mouse model. Neurotox Res, 21(4), 393–404. 10.1007/s12640-011-9294-3

34. Thakur, S., Dhapola, R., Sarma, P., Medhi, B., & Reddy, D. H. (2023). Neuroinflammation in Alzheimer’s Disease: Current Progress in Molecular Signaling and Therapeutics. Inflammation, 46(1), 1–17. 10.1007/s10753-022-01721-1

35. Vaine, C. A., Shin, D., Liu, C., Hendriks, W. T., Dhakal, J., Shin, K., Sharma, N., & Bragg, D. C. (2017). X-linked Dystonia-Parkinsonism patient cells exhibit altered signaling via nuclear factor-kappa B. Neurobiol Dis, 100, 108–118. 10.1016/j.nbd.2016.12.016

36. Westenberger, A., Reyes, C. J., Saranza, G., Dobricic, V., Hanssen, H., Domingo, A., Laabs, B. H., Schaake, S., Pozojevic, J., Rakovic, A., Grutz, K., Begemann, K., Walter, U., Dressler, D., Bauer, P., Rolfs, A., Munchau, A., Kaiser, F. J., Ozelius, L. J., Klein, C. (2019). A hexanucleotide repeat modifies expressivity of X-linked dystonia parkinsonism. Ann Neurol, 85(6), 812–822. 10.1002/ana.25488

37. Yang, H. S., Zhang, C., Carlyle, B. C., Zhen, S. Y., Trombetta, B. A., Schultz, A. P., Pruzin, J. J., Fitzpatrick, C. D., Yau, W. W., Kirn, D. R., Rentz, D. M., Arnold, S. E., Johnson, K. A., Sperling, R. A., Chhatwal, J. P., & Tanzi, R. E. (2022). Plasma IL-12/IFN-gamma axis predicts cognitive trajectories in cognitively unimpaired older adults. Alzheimers Dement, 18(4), 645–653. 10.1002/alz.12399

38. Yu, G., Liang, Y., Zheng, S., & Zhang, H. (2018). Inhibition of Myeloperoxidase by N-Acetyl Lysyltyrosylcysteine Amide Reduces Oxidative Stress-Mediated Inflammation, Neuronal Damage, and Neural Stem Cell Injury in a Murine Model of Stroke. J Pharmacol Exp Ther, 364(2), 311–322. 10.1124/jpet.117.245688

39. Yu, H., Liu, Y., Wang, M., Restrepo, R. J., Wang, D., Kalogeris, T. J., Neumann, W. L., Ford, D. A., & Korthuis, R. J. (2020). Myeloperoxidase instigates proinflammatory responses in a cecal ligation and puncture rat model of sepsis. Am J Physiol Heart Circ Physiol, 319(3), H705–H721. 10.1152/ajpheart.00440.2020

